# Low level of knowledge about COVID-19 among a sample of Deaf persons in Ghana

**DOI:** 10.1101/2022.06.17.22276229

**Authors:** Reginald Arthur-Mensah, Jacob Nartey Quao, Louisa Yeboah, Zanu Dassah, Abigail Agartha Kyei

**Affiliations:** Department of Nursing and Midwifery, Faculty of Health and Allied Sciences, Pentecost University, Accra, Ghana; Health Information Office, Ghana Health Service, Accra, Ghana; Sign Language Unit, Maamobi General Hospital, Accra, Ghana; Faculty of Education, Pentecost University, Accra, Ghana; Out Patient Department, Ga North Municipal Hospital, Accra, Ghana; Human Resource Health Directorate, Ghana Health Service, Accra, Ghana

**Author notes:** **Corresponding author**:; (RAM). These authors contributed equally to this work (Reginald Arthur-Mensah, Jacob Nartey Quao, Abigail Agartha Kyei). These authors also contributed equally to this work (Louisa Yeboah, Zanu Dassah).

**Keywords:** Attitude, Ayawaso North Municipality, COVID-19, Deaf persons, Ghana, Knowledge, Practice

## Abstract

Global observations have shown that the success or failure in preventing and controlling the spread of COVID-19 largely relies on human behaviours. Human behaviours in preventing and controlling the spread of the disease principally, is dependent on the level of knowledge of the disease, the attitudes adopted by persons due to the level of knowledge of the disease and the decision to adhere to the preventive practices (KAP) of the disease. Since the beginning of this pandemic, numerous studies have been conducted to investigate the KAP on the novel COVID-19 among diverse demographic groups. However, no reported studies have been found on the KAP of the COVID-19 pandemic among the deaf in various populations around the world. This study sought to assess the KAP of COVID-19 among deaf persons in the Greater Accra region of Ghana.

The design of this study utilized the knowledge, attitude and practice (KAP) survey. Good attitude and adherence to the preventive practices of COVID-19 was observed among the deaf persons. However, knowledge about the science of the disease was lacking. Educational campaigns about COVID-19 should also emphasize the teaching and understanding of the science of the virus and the disease to its audience.

## Introduction

The novel coronavirus disease 2019 (COVID-19) is the latest in the list of pandemics that is reforming every level of human survival. The pandemic is an infectious disease caused by the severe acute respiratory syndrome coronavirus 2 (SARS-CoV-2) virus [1]. The WHO declared the coronavirus disease outbreak a public health emergency of international concern on 30th January 2020, and a pandemic on 11th March 2020 [2].

Continuing studies have aligned the full-length genome sequence of SARS CoV-2 and other available genomes of beta coronaviruses. Results have shown that the closest relationship of SARS-CoV-2 is with the bat SARS-like coronavirus strain BatCov RaTG13, with an identity of 96%. These studies suggest that SARS-CoV-2 could be of bat origin, and SARS-CoV-2 might be naturally evolved from bat coronavirus RaTG13 [3,4]. Furthermore, [5,6] also showed beta coronavirus isolated from pangolins has a sequence similarity of up to 99% with the currently infecting human strain, suggesting the potential for pangolins to be the intermediate host. SARS CoV-2 belongs to the subfamily *Coronavirinae* in the family of Coronaviridae. The genome of CoVs is a single-stranded positive-sense ribose nucleic acid (+ssRNA) [7]. Generally, coronaviruses cause respiratory, digestive, and nervous system diseases in humans and many other animals [8].

The virus spreads through small liquid particles from an infected person’s mouth or nose when they cough, sneeze, speak, sing or breathe out. These liquid particles range from large respiratory droplets to small aerosols [9].

Anyone can get infected with the virus and become ill at any time. However, most people infected with the virus will experience mild to moderate respiratory illness and recover without requiring special treatment. Others will become seriously ill and require medical attention. Older people and those with underlying medical conditions like cardiovascular disease, diabetes, chronic respiratory disease, hypertension and cancer are more likely to develop serious illness [5,10,11,12].

Symptoms of the disease have been classified into three levels according to the severity of the disease; the less common symptoms, the most common symptoms and serious symptoms. The less common symptoms include experiencing sore throat, headaches, aches and pains, diarrhoea, noticing a rash on skin, or discoloration of fingers or toes red or irritated eyes. The most common symptoms include experiencing fever, tiredness, loss of taste or smell and having a dry cough. The serious symptoms include difficulty breathing or shortness of breath, loss of speech or mobility, or confusion and chest pains [13].

On average it takes 5 – 6 days from when someone is infected with the virus for symptoms to show up, however it can take up to 14 days for symptoms to show up. Important complications due to the infection include acute respiratory distress syndrome, sepsis, digestive stress, liver injury, hyperinflammatory response, multiorgan failure, thromboembolism and vascular damage [14,15].

All viruses, including SARS-CoV-2, mutate over time. Most changes have little to no impact on the virus’ properties. However, some changes may affect the virus’s properties, such as how easily it spreads, the associated disease severity, or the performance of vaccines, therapeutic medicines, diagnostic tools, or other public health and social measures [16]. The WHO, in collaboration with partners, expert networks, national authorities, specialist institutions and researchers have been monitoring and assessing the evolution of SARS-CoV-2 since January 2020. During late 2020, the emergence of variants that posed an increased risk to global public health prompted the characterization of variants, namely, Variants of Concern (VOCs), Variants of Interest (VOIs) and Variants under Monitoring (VUM) in order to prioritize global monitoring and research, and ultimately to inform the ongoing response to the COVID-19 pandemic.

VOC are case defined as variants that have been demonstrated to be associated with one or more of the following changes at a degree of global public health significance:

1. Increase in transmissibility or detrimental change in COVID-19 epidemiology; OR
2. Increase in virulence or change in clinical disease presentation; OR
3. Decrease in effectiveness of public health and social measures or available diagnostics, vaccines, therapeutics [16].

Currently circulating VOCs include the Delta and Omicron variant. Previously circulating VOCs include the Alpha, Beta and Gamma variants [16].

VOI are case defined as variants that have been demonstrated to be associated with one or more of the following changes at a degree of global public health significance:

1. Genetic changes that are predicted or known to affect virus characteristics such as transmissibility, disease severity, immune escape, diagnostic or therapeutic escape;
2. Identified to cause significant community transmission or multiple COVID-19 clusters, in multiple countries with increasing relative prevalence alongside increasing number of cases over time, or other apparent epidemiological impacts to suggest an emerging risk to global public health.

Circulating VOIs include Epsilon, Zeta, Eta, Theta, Iota, Kappa, Lambda and Mu [16]. VUMs are case defined as variant with genetic changes that are suspected to affect virus characteristics with some indication that it may pose a future risk, but evidence of phenotypic or epidemiological impact is currently unclear, requiring enhanced monitoring and repeat assessment pending new evidence. VUM include B.1.640 and XD [16].

Cardinal among the ways to prevent and control the infection is to be well informed about the disease and how the virus spreads. The following current precautionary measures are recommended according to public and social health standards;

1. Wearing properly face protection at all times e.g., properly fitting face mask
2. Practicing physical distancing (at least 1 meter apart) from others, even if they do not appear sick
3. Practicing good respiratory hygiene by covering the mouth and nose with a cloth when sneezing or coughing or sneezing or coughing in your flexed elbow in the absence of a cloth
4. Choosing open, well-ventilated spaces over closed ones
5. Washing the hands regularly with soap under running water for at least 30 seconds
6. Cleaning the hands intermittently with an alcohol-based hand sanitizer
7. Getting vaccinated
8. Staying home or indoors if you are experiencing any of the symptoms [16].

Although vaccination against COVID-19 is ongoing, no specific treatment is available for COVID-19, although effective therapeutic approaches are in development [17,18]. Global observations have shown that the success or failure in preventing and controlling the spread of the disease largely relies on human behaviours. Human behaviours in preventing and controlling the spread of the disease principally, is dependent on the level of knowledge of the disease, the attitudes adopted by persons due to the level of knowledge of the disease and the decision to adhere to the preventive practices (KAP) of the disease. Since the beginning of this pandemic, numerous studies have been conducted to investigate the KAP on the novel COVID-19 among diverse demographic groups. These studies have allowed us to identify groups that have adequate or deficient knowledge to the disease, groups that have developed positive or negative attitudes to the disease and groups that are adhering to or otherwise to the preventive practices of the disease. However, no reported studies have been found on the KAP of the COVID-19 pandemic among the deaf in various populations around the world. Comprehensive understanding on the life cycle of SARS-CoV-2 and the interaction of the virus with hosts are necessarily important in the fight against the COVID-19 pandemic. As well, to facilitate successful and effective management of the COVID-19 pandemic globally, all-inclusive awareness of the KAP of disease are fundamentally important. It is essential to understand individuals’ knowledge, attitudes and adherence to practices toward COVID-19 since public adherence to health guidelines relies heavily on these facets. KAP of COVID-19 among all majority and minority demographic groups ought to be assessed. This study sought to assess the KAP of COVID-19 among a deaf community in the Greater Accra region of Ghana.

## Materials and Methods

### Research design

The design of this study utilized the knowledge, attitude and practice (KAP) survey. KAP surveys contain a set of questions segmented into knowledge, attitude and practices of a particular phenomenon of interest to receive responses to these questions. Responses to KAP surveys provides access to quantitative or qualitative information for the purpose of the KAP survey. Firstly, KAP surveys provides information about what is known, believed and done about phenomenon of interest among a target population. It then reveals misunderstandings, misconceptions and misbehaviors that signify hindrances to the phenomenon of interest. It further exposes aspects of the phenomenon of interest that are not yet known and practiced among a target population [19].

In the current study, the KAP survey will help us to measure the KAP of COVID-19 and to provide new tangents of the pandemic’s reality among a deaf community in Ghana. The results from the survey will establish novel baseline information on the KAP of COVID-19 among the deaf community for use in future assessments. It will further help to justify the effectiveness or otherwise of the public health interventions against COVID-19 towards health-related behaviour change among the populace.

### Study area

The Deaf community in the Ayawaso North Municipality were sampled for this study. The Ayawaso North Municipality is one of the 29 Metropolitan, Municipal and District Assemblies (MMDAs) in the Greater Accra region of Ghana. The topography of the area, climate, vegetation, housing structures, road networks, ethnic and cultural diversity, occupation of persons and religious affiliation is similar to any municipality of the Greater Accra Region.

### Study sample

Deaf persons in the Ayawaso North Municipality were sampled for the study. They included both male and female deaf persons above the age of ten years who could communicate the American/Ghanaian Sign Language (A/GSL).

### Sample size and sampling procedure

The census method of sampling was used to include all deaf persons at the Ayawaso North Municipality into the study. Census sampling is a statistical method of sampling that studies all the members of a population in a research study. Results obtained from census samples are unabridged, useful and directly prognostic [20]. In all, 144 deaf persons were sampled in the current study.

### Data collection instrument

The KAP COVID-19 instrument developed by [21] was adapted for this study. Our KAP COVID-19 questionnaire consisted of 12 question items on knowledge, 6 question items on attitudes and 9 question items on adherence to preventive practices of COVID-19. Responses were measured on a numeric rating scale. Question items on knowledge were rated as I do not know = 0 and Yes = 1. Question items on attitudes were rated as No = 0, I am not sure = 1 and Yes = 2. Question items on adherence to preventive practices of COVID-19 were rated as Never = 0, Sometimes = 1, Often = 2 and Always = 3. The psychometric properties, validity and reliability of the KAP COVID-19 questionnaire have been reported [21].

### Data collection procedure

Data collection was scheduled during the second round of the National COVID-19 mass vaccination at the Municipality. These second round of vaccinations was to help achieve herd immunity against the virus in the country. A cubicle for our data collection was setup alongside the National COVID-19 mass vaccination team setup during the second round of vaccination. As deaf persons came to the center to be registered for the vaccination, we collected our data before they received the jab. In the cubicle, they were warmly welcomed, given a seat and served with water. The purpose of the study was then thoroughly explained to deaf persons. Afterwards, they were given opportunity to ask questions and seek clarification on any issue relating to the study. Those who agreed to take part in the study, signed or thumb printed the consent forms before data was collected. Some questionnaires were self-administered whilst others were completed with the help of the researchers.

Notably, data was collected by researchers who were familiar with the deaf culture, thus, all values peculiar to the deaf culture were observed. All communication was done using the A/GSL. Additionally, COVID-19 protocols were observed between the researchers and the deaf persons during the period of data collection. After data collection, deaf persons proceeded to take their second jab of COVID-19 vaccine. Data was collected from April 19th to 30th, 2022.

### Ethical considerations

Ethical approval to conduct this study was given by the Ghana Health Service (GHS) Institutional Review Board (IRB) after the study protocol was reviewed. Additionally, permission to conduct the study among the deaf persons in the municipality was given by the Municipal Health Director, the Municipal Public Health Nurse, the Municipal Disease Control Officer, the Municipal Health Promotion Officer, the Municipal Health Information Officer, the Municipal Chief Director, the Municipal Social Welfare Director and the head of the Ghana National Association of the Deaf representatives in the district. Study details were thoroughly explained to our sample before data collection. Our sample were given opportunity to ask questions and seek clarification on any issue relating to the study before data collection. This was done to prevent issues of dishonesty. Those who agreed to take part in the study signed or thumb printed the consent forms before data was collected. Some questionnaires were self-administered whilst others were completed with the help of the researchers. Additionally, the methods for the study ensured that there was no harm in any way to the sample during data collection and they were assured of confidentiality. They were also informed of the voluntary nature of the study participation or withdrawal without any negative consequences. The study also ensured that privacy of the sample was respected and no act compromised their privacy. For deaf persons who were under 18 years, written informed consent was obtained from either of their parents/guardians before data collection. They were also taken through the ethical procedures outlined above.

## Data analysis

Data analysis was performed using IBM® SPSS software version 23.0 (SPSS Inc, Chicago, IL, USA). Firstly, frequencies and percentages were used to present the demographic details of our sample. Secondly, frequencies and percentages were used to present the responses to the knowledge and attitude subscales. Average scores using modal values were used to present the results of the adherence to the preventive practices of COVID-19 subscale.

## Results

### Demographic details of Deaf persons

A total of 144 deaf persons were sampled in the study. This number reflected all deaf persons who had registered with the Municipal Directorate. They comprised 68/144 (47.2%) males and 76/144 (52.8%) females. Their mean age was 36 years (±12.84). The range of their ages was from 12 years to 65 years. Regarding their educational attainment, the majority, 48/144 (33.3) attained up to Junior High School level of education. 28/144 (19.4%) had no formal education. About a quarter of the deaf persons, 26/144 (18.1%) had attained up to the Basic level and the Senior High School level of education and 16/144 (11.1%) had attained up to the tertiary level of education. The bulk, 64/144 (44.4%) of the deaf persons were unemployed, 40/144 (27.8%) were doing their own businesses, 24/144 (16.7%) were working at various private sector businesses and 16/144 (11.1%) were Government workers. 116/144 (80.6%) of deaf persons were unmarried and the rest, 28/144 (19.4%) were married (Table 1).

**Table 1:** Demographic details of Deaf persons.

| **Demographic details** | ***f* (%)** |
| --- | --- |
| **Gender** |  |
| Males | 68 (47.2) |
| Females | 76 (52.8) |
| **Total** | 144 (100) |
| **Educational level** |  |
| No formal education | 28 (19.4) |
| Primary | 26 (18.1) |
| Junior High School | 48 (33.3) |
| Senior High School | 26 (18.1) |
| Tertiary | 16 (11.1) |
| **Total** | 144 (100) |
| **Occupation** |  |
| Unemployed | 64 (44.4) |
| Government worker | 16 (11.1) |
| Private business employee | 24 (16.7) |
| Doing own business | 40 (27.8) |
| **Total** | 144 (100) |
| **Marital status** |  |
| Married | 28 (19.4) |
| Unmarried | 116 (80.6) |
| **Total** | 144 (100) |

### Level of knowledge of COVID-19 among Deaf persons

Twelve question items were asked the deaf persons regarding the knowledge of COVID-19 subscale. Responses to the questions items were rated on a numeric scale as I do not know = 0 and Yes = 1. Results showed that majority of the deaf persons were not knowledgeable on 8 questions items. This represented a lack of knowledge of 67% of the items of the knowledge subscale (Table 2).

**Table 2:** Level of knowledge of COVID-19 among Deaf persons.

| Question items | Response option | *f* (%) |
| --- | --- | --- |
| **1. COVID-19 is a disease caused by a virus** | **I do not know** | **124 (86.1)** |
|  | Yes | 20 (13.9) |
|  | Total | 144 (100) |
| **2. The virus is an RNA virus** | **I do not know** | **138 (95.8)** |
|  | Yes | 6 (4.2) |
|  | Total | 144 (100) |
| **3. The virus is called SARS COV-2** | **I do not know** | **140 (97.2)** |
|  | Yes | 4 (2.8) |
|  | Total | 144 (100) |
| **4. On average, it takes 5 – 14 days when someone is infected with the virus for symptoms to show up** | **I do not know** | **130 (90.3)** |
|  | Yes | 14 (9.7) |
|  | Total | 144 (100) |
| **5. COVID-19 is currently being diagnosed using antibodies, proteins and gene detection methods** | **I do not know** | **140 (97.2)** |
|  | Yes | 4 (2.8) |
|  | Total | 144 (100) |
| **6. Current treatment for COVID-19 is symptomatic treatment** | **I do not know** | **90 (62.5)** |
|  | Yes | 54 (37.5) |
|  | Total | 144 (100) |
| **7. To this date, there is no antiviral agent specific to the virus.** | **I do not know** | **122 (84.7)** |
|  | Yes | 22 (15.3) |
|  | Total | 144 (100) |
| **8. The virus keeps mutating** | **I do not know** | **94 (65.3)** |
|  | Yes | 50 (34.7) |
|  | Total | 144 (100) |
| **9. COVID-19 is spread through droplet transmission** | **I do not know** | **46 (31.9)** |
|  | Yes | 98 (68.1) |
|  | Total | 144 (100) |
| **10. Anyone can get infected with the virus** | **I do not know** | **22 (15.3)** |
|  | Yes | 122 (84.7) |
|  | Total | 144 (100) |
| **11. The main clinical symptoms of COVID-19 include fever, dry cough, tiredness, loss of taste and smell** | I do not know | 24 (16.7) |
|  | Yes | 120 (83.3) |
|  | Total | 144 (100) |
| **12. Persons with underlying health conditions might suffer serious illness of the disease.** | I do not know | 60 (41.7) |
|  | Yes | 84 (58.3) |
|  | Total | 144 (100) |

### Attitude of Deaf persons towards COVID-19

Six question items were asked concerning the attitude of deaf persons towards COVID-19 subscale. Responses to the questions items were rated on a numeric scale as No = 0, I am not sure = 1 and Yes = 2. Results showed that majority of the deaf persons had good attitude towards COVID-19. Deaf persons showed good attitude in all 6 items of the attitude subscale. As a point worthy of note, all deaf persons said “yes” to getting vaccinated against COVID-19 on the day of data collection and any other time and were ready to encourage their family and friends to get vaccinated against COVID-19 (Table 3).

**Table 3:**
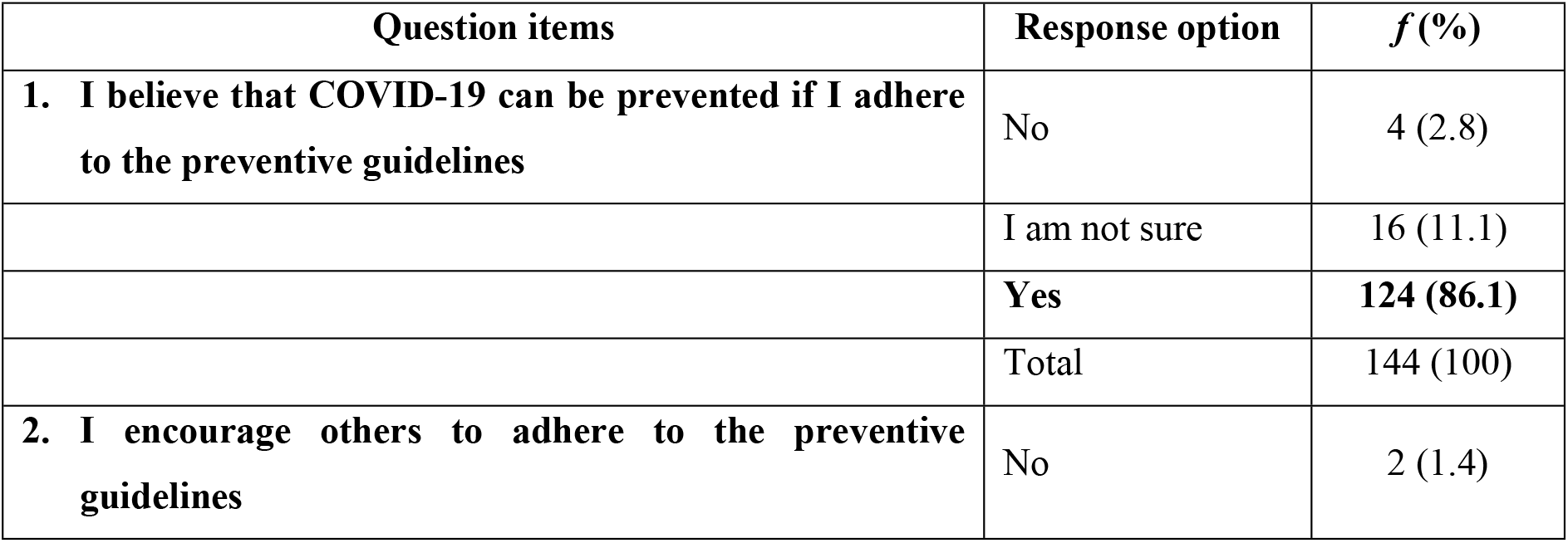
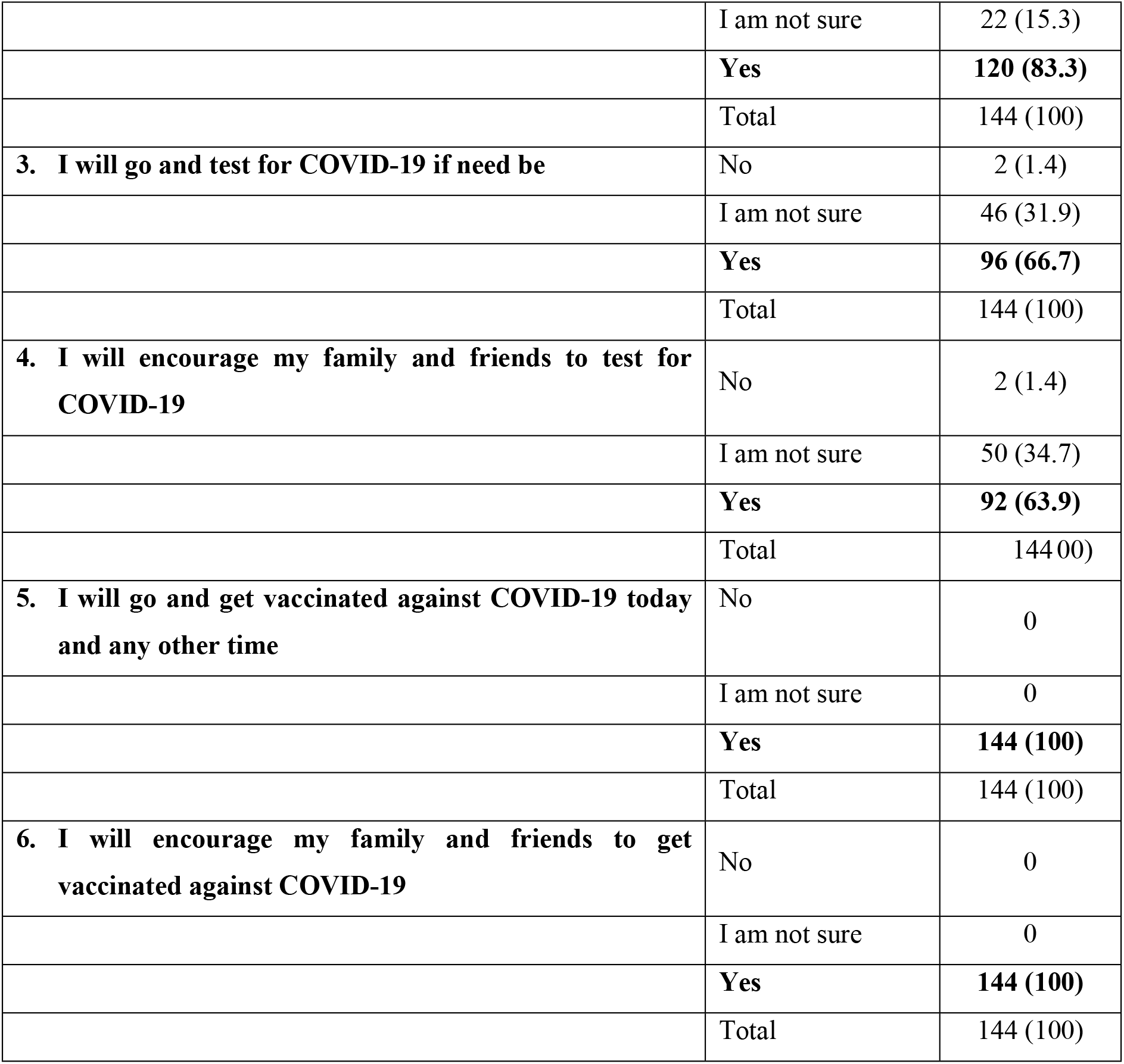
Attitude of Deaf persons towards COVID-19.

### Adherence to preventive practices of COVID-19 among Deaf persons

Nine question items were asked the deaf persons pertaining to their adherence to preventive practices of COVID-19 subscale. Responses to the questions items were rated on a numeric scale as Never = 0, Sometimes = 1, Often = 2 and Always = 3. Average score using modal values were used to represent the aggregate responses of our sample. Results showed that deaf persons “always” practised 5 items in the adherence to preventive practices subscale and “sometimes” practised 4 items in the subscale. This represented a strict adherence to the preventive practices of COVID-19 of 56% of the items of the adherence to COVID-19 preventive practices subscale (Table 4).

**Table 4:** Adherence to preventive practices of COVID-19 among Deaf persons.

| Question items | Modal score |
| --- | --- |
| 1. I wear face protection at all times | 3.00 |
| 2. I avoid physical contact (e.g., handshakes, hugs, etc.) with others as much as possible | 3.00 |
| 3. I choose open, well-ventilated spaces over closed ones | 3.00 |
| 4. I wash my hands regularly with soap under running water for at least 30 seconds | 3.00 |
| 5. I clean my hands intermittently with an alcohol-based hand sanitizer | 3.00 |
| 6. I practice physical distancing (at least 1 meter apart) from others, even if they do not appear sick as much as possible | 1.00 |
| 7. I practicing good respiratory hygiene by covering my mouth and nose with a cloth when sneezing or coughing or I sneeze or cough in my flexed elbow in the absence of a cloth | 1.00 |
| 8. I avoid touching my eyes, nose and mouth as much as possible | 1.00 |
| 9. I stay home or indoors if I am experiencing any of the symptoms and any other symptoms, I am not sure of | 1.00 |

## Discussion

Having observed with ardent interest in the scientific literature on the scope of the COVID-19 disease since its inception, specifically on the knowledge, attitude and adherence to the preventive practices (KAP) of the disease, we noticed few and unmethodical reports on the KAP of COVID-19 among deaf persons globally. We therefore set forth to contribute to the scientific literature on the scope of COVID-19 on the KAP of COVID-19 among deaf persons in Ghana from a sample of deaf persons in the Ayawaso North Municipality.

Good attitude and adherence to the preventive practices of COVID-19 was observed among the deaf persons. However, knowledge about the science of the disease was lacking. The good attitude and adherence to the preventive practices of COVID-19 observed reflects the mass public health preventive campaigns to COVID-19 by various stakeholders including the Ministry of Health (MOH), Ghana Health Service (GHS), Ministry of Information (MOI), media houses and individuals about the preventive measures of the disease in Ghana. All campaigns about the disease including the periodic ‘presidential updates on measures taken in the fight against the pandemic in Ghana’, largely focuses on the preventive practices against the disease. Besides, the GHS through it Municipal Health Directorates (MHD) routinely organized targeted public health prevention campaigns against COVID-19 for deaf persons registered with the municipalities. In these campaigns, deaf persons were constantly updated on aseptic ways to handle, wear and dispose of their facemasks, the right steps to wash their hands, the right steps and moments to use hand sanitizers, how to practice physical distancing, how to practice good respiratory hygiene and to get vaccinated when the vaccines arrive in the country and they are called upon to get vaccinated.

As is the case, targeted special public health education about the COVID-19 vaccination was done by GHS through its MHDs for all deaf persons registered with the municipalities when the vaccines were received in the country and it was time to vaccinate the populace. It was during one of such special interventions for deaf persons, i.e., the nationwide second dose of COVID-19 vaccination for deaf persons, that this study was conducted. Additionally, very early in the pandemic, during televised live briefings and recorded sessions about the situation of the pandemic in the country by the President or the Minister for Information or the Minister for Health or the Director of the GHS, deaf communities were supported by having sign language interpreters during these broadcast. These broadcasts also never ended without reiterating the adherence to the preventive practices against the disease.

Regarding the knowledge of the science of COVID-19, the municipal health directorate were shown the findings of this study. They agreed to it, realizing their primal goal has been on teaching deaf persons on how to protect themselves from the disease rather than the science of the disease. Deficient knowledge was found on the causative agent of the disease, the nature and dynamics of the virus, the name of the virus, the incubation period of the virus and the detection and diagnostic means of the virus. This is disturbing because these items represent the basic information about the pandemic. One of the major concerns about the pandemic was ensuring access to right information about the virus and the disease to all persons. Concerning deaf persons, many communication gaps sort to be bridged by various advocacy institutions for deaf persons working in partnership in this period. One of such interventions was the development of new vocabulary for COVID-19 terminology for deaf persons [22]. This helped the deaf community and sign language interpreters engaged in various formal and informal interpretation of COVID-19 related information resources. Yet still, this study shows that gaps still persist. A relational barrier may be seen here. On the part of the sign language interpreters, it may be likely that interpreters were not keeping themselves abreast with the international sign language for COVID-19 or interpreters were unable to break down the scientific terminologies e.g., SARS COV-2, RNA, droplet infection, underlying health conditions, antibodies, proteins and gene detection methods, mutation, herd immunity etc. associated with the disease to the deaf persons. On the part of deaf persons too, it is probable that deaf persons did not understand or were unfamiliar with the scientific terminologies associated with the disease. In such situations, interpreters must use a scenario to describe what they are communicating or finger spell what they are describing to deaf persons. The low level of education recorded amongst deaf persons could influence their appreciation of these scientific terms or rather simply, deaf persons were also not keeping themselves informed and updated on the progress of the disease. Apropos, getting knowledge about the disease was passive among deaf persons.

The knowledge of the disease, the attitudes adopted by persons due to the level of knowledge of the disease and the decision to adhere to the preventive practices (KAP) of the disease is vital in the control and management of the disease in Ghana and globally.

## Conclusions

The glaring challenges to social inclusion posed by the pandemic was examined in terms of the KAP of COVID-19 among a deaf community in Ghana. Regarding knowledge of COVID-19, findings from the study showed that majority of the deaf persons were not knowledgeable on 8 questions items. This represented a lack of knowledge of 67% of the items of the knowledge subscale. Concerning attitude to COVID-19, findings showed that majority of the deaf persons had good attitude towards COVID-19. Deaf persons showed good attitude in all 6 items of the attitude subscale. Pertaining to the adherence to the preventive practices against the disease, findings showed that deaf persons “always” practised 5 items in the adherence to preventive practices subscale and “sometimes” practised 4 items in the subscale. This represented a strict adherence to the preventive practices of COVID-19 of 56% of the items of the adherence to preventive practices subscale.

### Recommendations

Educational campaigns about COVID-19 should also emphasize the teaching and understanding of the science of the virus and the disease to its audience. New concepts and vocabulary especially relating to health concepts and vocabulary should be published far and wide regularly to keep the deaf community and sign language interpreters updated, to ensure efficient communication. Health institutions should continually employ the services of qualified sign language interpreters to guarantee the incessant inclusion of deaf persons in health activities.

## Data Availability

Data regarding this work is available from the corresponding author upon reasonable request.

https://17026/dans-x3n-pn69

## Acknowledgements

We are grateful to the Municipal Health Director, the Municipal Public Health Nurse, the Municipal Disease Control Officer, the Municipal Health Promotion Officer, the Municipal Health Information Officer, the Municipal Chief Director, the Municipal Social Welfare Director and the head of the Ghana National Association of the Deaf representatives of the Ayawaso North Municipality for allowing us to conduct this important study. We are indebted to the deaf persons for sharing their time with us.

## Notes

### Competing Interest Statement

The authors have declared no competing interest.

### Funding Statement

The authors received no specific funding for this work.

### Author Declarations

Ethical approval to conduct this study was given by the Ghana Health Service (GHS) Institutional Review Board (IRB) after the study protocol was reviewed.

